# Use of High Flow Nasal Therapy to Treat Moderate to Severe Hypoxemic Respiratory Failure in COVID-19

**DOI:** 10.1101/2020.05.22.20109355

**Authors:** Maulin Patel, Andrew Gangemi, Robert Marron, Junad Chowdhury, Ibraheem Yousef, Matthew Zheng, Nicole Mills, Lauren Tragesser, Julie Giurintano, Rohit Gupta, Matthew Gordon, Parth Rali, Gilbert D’Alonzo, David Fleece, Huaqing Zhao, Nicole Patlakh, Gerard J. Criner, for the Temple University COVID-19 Research Group

## Abstract

Invasive mechanical has been associated with high mortality in COVID-19. Alternative therapy of High flow nasal therapy (HFNT) has been greatly debated around the world for use in COVID-19 pandemic due to concern for increased healthcare worker transmission.

**Methods:** This was a retrospective analysis of consecutive patients admitted to Temple University Hospital in Philadelphia, Pennsylvania, from March 10, 2020, to April 24, 2020 with moderate to severe respiratory failure treated with High Flow nasal therapy (HFNT). Primary outcome was prevention of intubation.

**Results:** Of the 445 patients with COVID-19, 104 met our inclusion criteria. The average age was 60.66 (±13.50) years, 49 (47.12 %) were female, 53 (50.96%) were African American, 23 (22.12%) Hispanic. Forty-three patients (43.43%) were smokers. SF and chest Xray scores had a statistically significant improvement from day 1 to day 7. 67 of 104 (64.42%) were able to avoid invasive mechanical ventilation in our cohort. Incidence of hospital/ventilator associated pneumonia was 2.9%. Overall, mortality was 14.44% (n=15) in our cohort with 13 (34.4%) in the progressed to intubation group and 2 (2.9%) in the non-intubation group. Mortality and incidence of VAP/HAP was statistically higher in the progressed to intubation group.

**Conclusion:** HFNT use is associated with a reduction in the rate of Invasive mechanical ventilation and overall mortality in patients with COVID-19 infection.

**Key Points:** *Key Question:* What is the utility of High Flow Nasal Therapy (HFNT) in COVID-19 related Hypoxemic Respiratory Failure?

*Bottom Line:* In this retrospective analysis of moderate to severe hypoxic respiratory failure for COVID 19 patients, 67 patients (65.4%) were able to avoid intubation despite severely low SF ratio (Mean 121.9).

*Why Read on:* HFNT has a significant role in COVID-19 for reducing rate of intubations and associated mortality

## Introduction

In December of 2019 a cluster of acute respiratory illnesses occurred in Hubei province, China, now known to be caused by a novel Coronavirus, also known as severe acute respiratory syndrome-coronavirus-2 (SARS-CoV-2). It has spread globally since with more than 2 million cases reported as of April 2020(1), (2). Severe hypoxemic respiratory failure is by far the most common reason for admission to intensive care units due to Coronavirus disease 2019 (COVID-19). In a report from Lombardi, Italy, of 1591 critically ill COVID-19 patients, 99% required respiratory support of at least supplemental oxygen and 88% (or 1150 patients) required invasive ventilation.(3) Another retrospective review of Wuhan hospitalized patients, including non-COVID-19 patients, showed 52% required respiratory support, of which 55% needed mechanical ventilation (4). Mortality of COVID-19 patients on invasive ventilation has been reported to be greater than 90% in Italy, China and New York. (3-5)

High flow nasal therapy (HFNT) is a non-invasive oxygen delivery system that allows for administration of humidified air-oxygen blends as high as 60 liters per minute and a titratable fraction of inspired oxygen as high as 100%. HFNT has shown effectiveness in other severe viral respiratory illnesses like influenza A and H1N1(6). Use of HFNT has led to lower progression to invasive ventilation compared to other forms of noninvasive oxygen therapy (7-9). By decreasing the incidence of invasive ventilation, HFNT has the potential advantage of theoretically decreasing the incidence of ventilator-associated pneumonia (VAP), as well as reduction in hospital resources which can be critical during times of increasing strain on the healthcare system. When compared with noninvasive positive pressure ventilation (NPPV), the use of HFNT is associated with similar rates of reintubation due to post-extubation respiratory failure. (10) However, no short-term mortality benefit has been reported using HFNT to treat acute hypoxemic respiratory failure. (7, 11, 12).

The Surviving Sepsis Guidelines for COVID-19 recommends using HFNT in patients with acute hypoxemic respiratory failure due to COVID-19 (13). However, others recommend against using HFNT fearing that it will create aerosolization of the COVID-19 virus and increase transmission to healthcare providers (14-16). In the few case series that report HFNT use in COVID-19 patients, its usage has ranged from 4.8 - 63.5% (17-20). In a recent report of patients who succumbed to COVID-19 in China, 34.5% were placed on HFNT alone; the authors postulated that use of HFNT may have contributed to a delay in intubation thereby increasing mortality (21).

Herein we present a retrospective analysis of the outcomes of COVID-19 patients with moderate-to-severe hypoxemic respiratory failure receiving HFNT at our center.

## Methods

The study was approved by the Temple University Institutional Review Board (TUIRB protocol number: 27051). A waiver of consent was granted due to the acknowledged minimal risk to the patients.

### Design

This was a retrospective analysis of consecutive patients admitted to Temple University Hospital in Philadelphia, Pennsylvania, from March 10, 2020, to April 24, 2020, for moderate to severe hypoxemia due to highly suspected or proven COVID-19 infection. Patients who presented to our hospital with fever or acute respiratory symptoms of unknown etiology were screened for COVID-19 infection. Patients included in analysis were those that tested positive for COVID-19 using nasopharyngeal real time reverse transcriptase PCR (RT-PCR) or patients with high clinical suspicion and findings suggestive of COVID-19 based on high-resolution computerized tomography (HRCT) of the chest (typical peripheral nodular or ground glass opacities without alternative cause (22, 23) with typical inflammatory biomarker profile.

Data including demographics, age, sex, comorbidities, body mass index (BMI), smoking status (current smoker, non-smoker), admission laboratory data including complete blood count (CBC) with differential, ferritin, lactate dehydrogenase (LDH), d-dimer, and C-reactive protein (CRP), treatments offered were collected for all of these patients. We also collected oxygen saturation to fraction of inspired oxygen ratio (SF ratio) on day of HFNT initiation, at day 7 after HFNT initiation or at discharge, whichever came earlier. SF was used as a surrogate for PF ratio (partial pressure of Oxygen/fraction of inspired Oxygen) as they have been correlated well in clinical trials (24).

### Radiology

Chest Radiographs (CXR) were graded by senior pulmonary and critical care fellows according to the RALES grading system (see Figure 1) previously studied in ARDS and organ donors (25). Chest X-rays were graded on the day of initiation of HFNT and earlier of discharge day or day 7.

**Figure 1:**
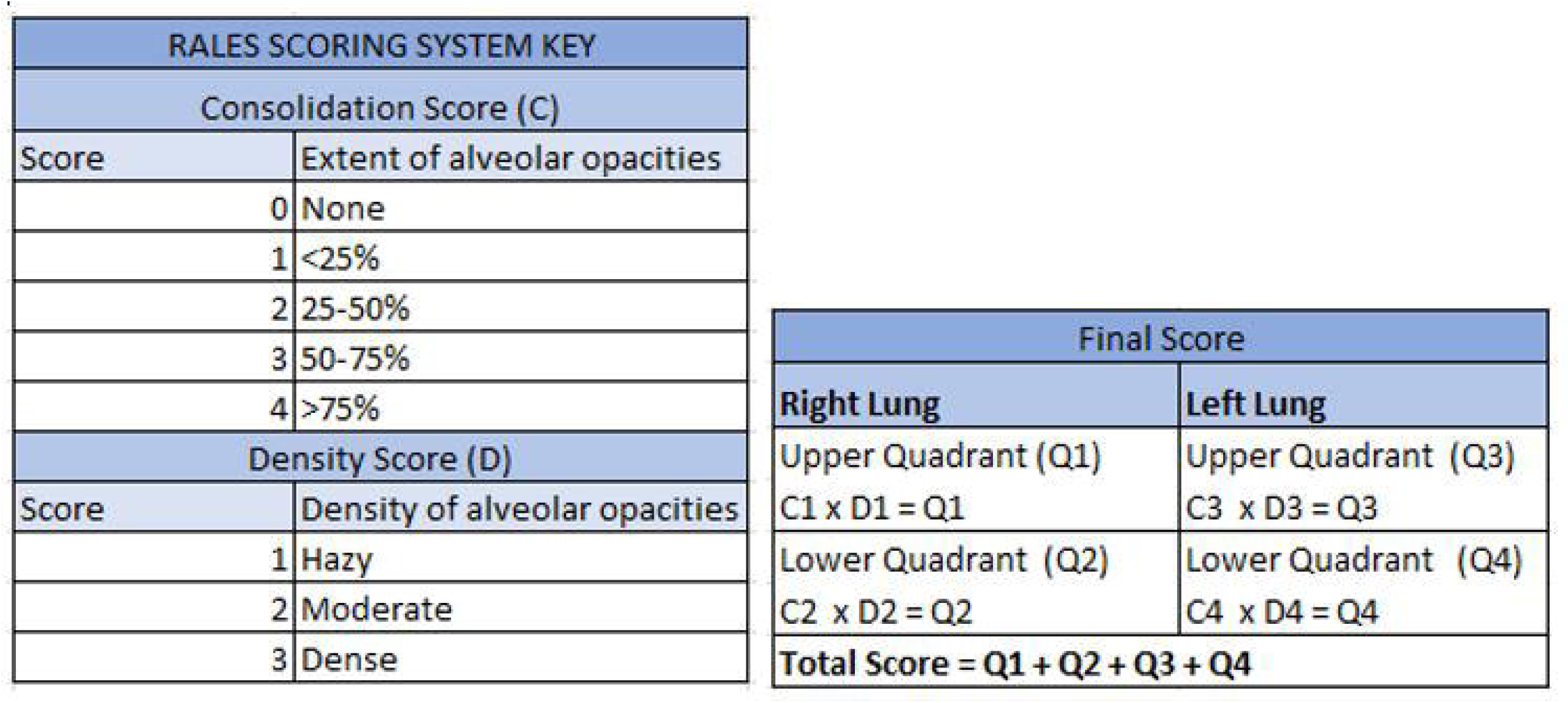
Radiographic Assessment of Lung Edema (RALE) grading system for Chest X Ray

### Respiratory therapy

All patients included in the analysis had moderate to severe hypoxemic respiratory failure and were on oxygen delivery via HFNT during the hospital course. Receipt of any other form of respiratory support initially was considered as exclusion criteria for the study. HFNT was provided with a humidified air-oxygen blend starting at 35 lpm with the fraction of inspired oxygen (F_I_O_2_) adjusted to maintain oxygen saturations ≥ 94%; further adjustments were made based on patients’ tolerance and goals of oxygenation. The initial temperature for the high flow setup was 37°C and was titrated between 34-37°C for patient comfort. Data on initial oxygenation support were collected which included the flow of air-oxygen blend in liters per minute and fractional percentage of inspired oxygen.

### Outcomes

Our primary outcome was the prevention of invasive mechanical ventilation (IMV) (%) with use of HFNT. Our secondary outcomes were mortality, change in oxygen saturation to fraction of inspired oxygen ratio (S-F ratio), change in RALE score of CXR, hospital length of stay (LOS) and hospital/ventilator acquired pneumonia. Hospital and ventilator acquired pneumonia was defined based on the presence of sputum positivity and treatment with antibiotics. Changes in S-F ratio were calculated by difference between S-F ratio at day 7 or discharge (whichever was earlier) versus day 1.

HFNT patients were divided into two groups: 1) progression to IMV (i.e., intubation group) and 2) continued HFNT support (i.e., non-intubation group). Patients who required NPPV are reported in the non-intubation group. Comparison was made between demographics, baseline laboratory values, and outcomes within the two groups. Improvements/worsening in oxygenation at day 7 and change in clinical parameters of heart (HR) and respiratory rates (RR) were also analyzed.

We constructed a prediction model for intubation for our cohort. All comorbidities, demographics, clinical and laboratory data were used to investigate parameters that could predict need for intubation. A cumulative comorbidity score (1 point allocated for each of the 5 comorbidities reported) and cumulative inflammatory laboratory marker score (1 point for each abnormal lab) were tested as predictors of intubation.

### Statistical methods

Continuous variables are presented as means (± standard deviation), and categorical variables as numbers and Frequency (percentages). Continuous variables were compared with the use of the two-sample t-test or paired t-test for categorical variables with the use of the Pearson chi-square test. Laboratory data were nonparametric and compared using Wilcox Rank-Sum test. Kaplan-Meier analysis was estimated for survival and compared by log-rank test.

To build a predictive model of the intubation, multivariable logistic regression was performed to determine the adjusted associations of the variables with intubation. The initial model included all the variables associated with intubation in univariate analyses for p<0.1. The final model that optimized the balance of the fewest variables with good predictive performance. Assessment of model performance was based on discrimination and calibration. Discrimination was evaluated using the C-statistic, which represents the area under the receiver operating characteristic (ROC) curve, where higher values represent better discrimination. Calibration was assessed by the Hosmer-Lemeshow test, where a p-value greater than 0.05 indicates adequate calibration.

All statistical tests were two-tailed, and P values of less than 0.05 were considered to indicate statistical significance. All statistical analyses were performed with the use of Stata 14.0 (StataCorp LP, College Station, TX).

## Results

### Patient population

894 patients admitted to Temple University Hospital between March 10, 2020, and April 24, 2020 who had suspected COVID-19 infection were retrospectively screened for our study. 445 patients had tested positive for COVID-19 by nasopharyngeal RT-PCR or were treated for high clinical suspicion based on typical CT imaging and inflammatory biomarker profile.

Of the 445 patients, 353 patients had hypoxemic respiratory failure requiring some form of oxygen therapy. The level of oxygen ranged from 2 L/min of oxygen via simple Nasal cannula to requiring invasive mechanical ventilation requiring positive pressure and 100% oxygen. 104 (23.3% of all COVID-19 positive patients) met our inclusion criteria of having moderate to severe COVID-19 related hypoxemic respiratory failure and were treated with HFNT (figure 2). The reported hypoxemia was moderate to severe with mean S-F ratio of 121.9 (range 79-225). Higher Chest Xray RALE scores were associated with more severe S-F ratios.

**Figure 2:**
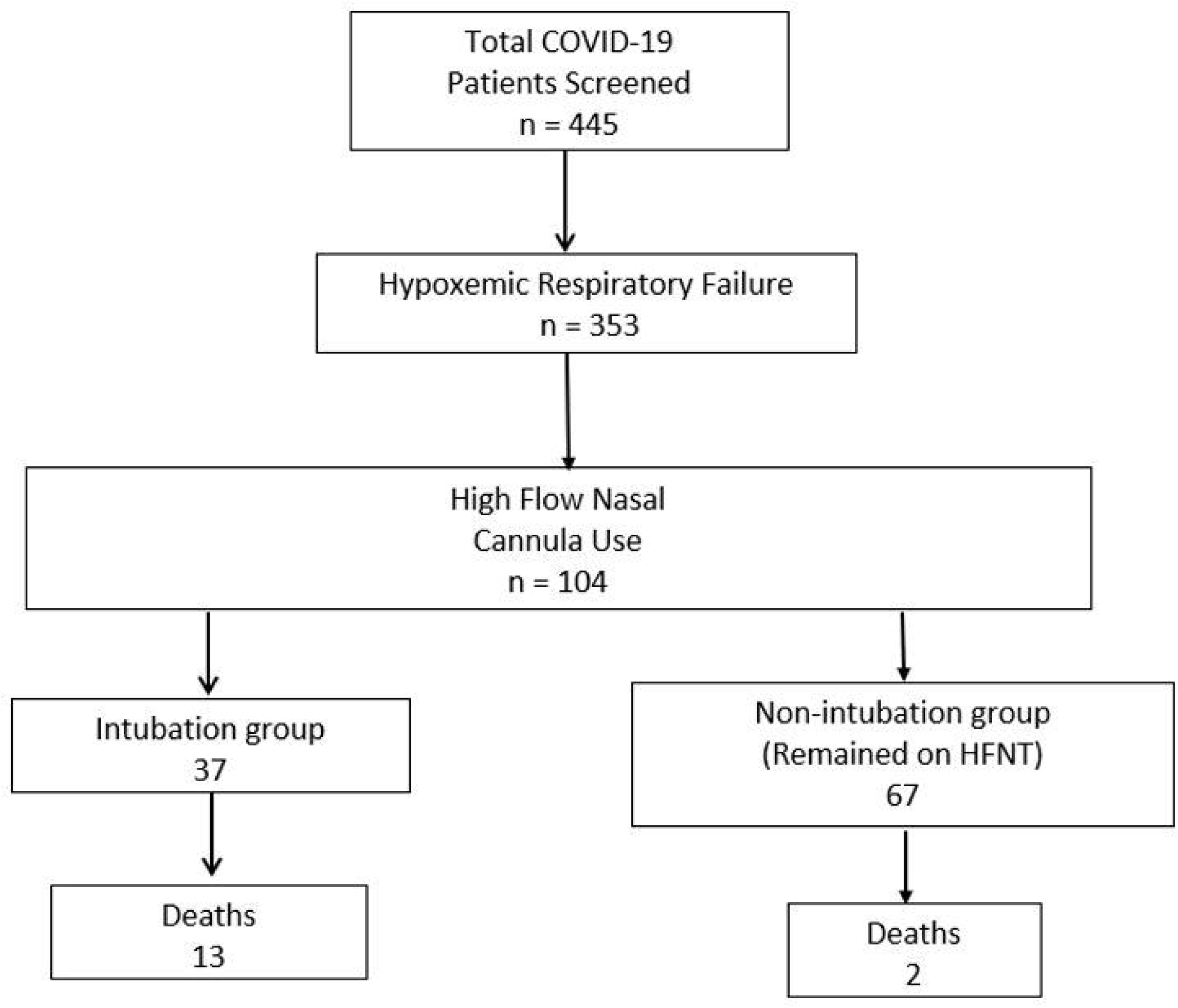
Flow chart demonstrating screening for our patients

The average age was 60.66 (±13.50) years, 49 (47.12 %) were female, 53 (50.96%) were African American, 23 (22.12%) Hispanic. Forty-three patients (43.43%) were smokers. The major comorbidities reported (in descending incidence) were hypertension, diabetes, lung disease, heart disease and chronic kidney disease (CKD) (Table 1). Nine (9.78%) patients were also on hemodialysis. Baseline S-F ratios were severely low at 121.9, corresponding to a P-F ratio of ~100. Elevated inflammatory markers (i.e., ferritin, CRP, D-dimer, fibrinogen, LDH, IL-6), creatinine along with transaminitis and lymphopenia were observed in all patients. In terms of treatments, azithromycin (57.2%) and steroids (64.71%) were the most frequently used therapies. Immunomodulators like sarilumab, anakinra, IVIG and tocilizumab were the next most commonly used therapies.

**Table 1:**
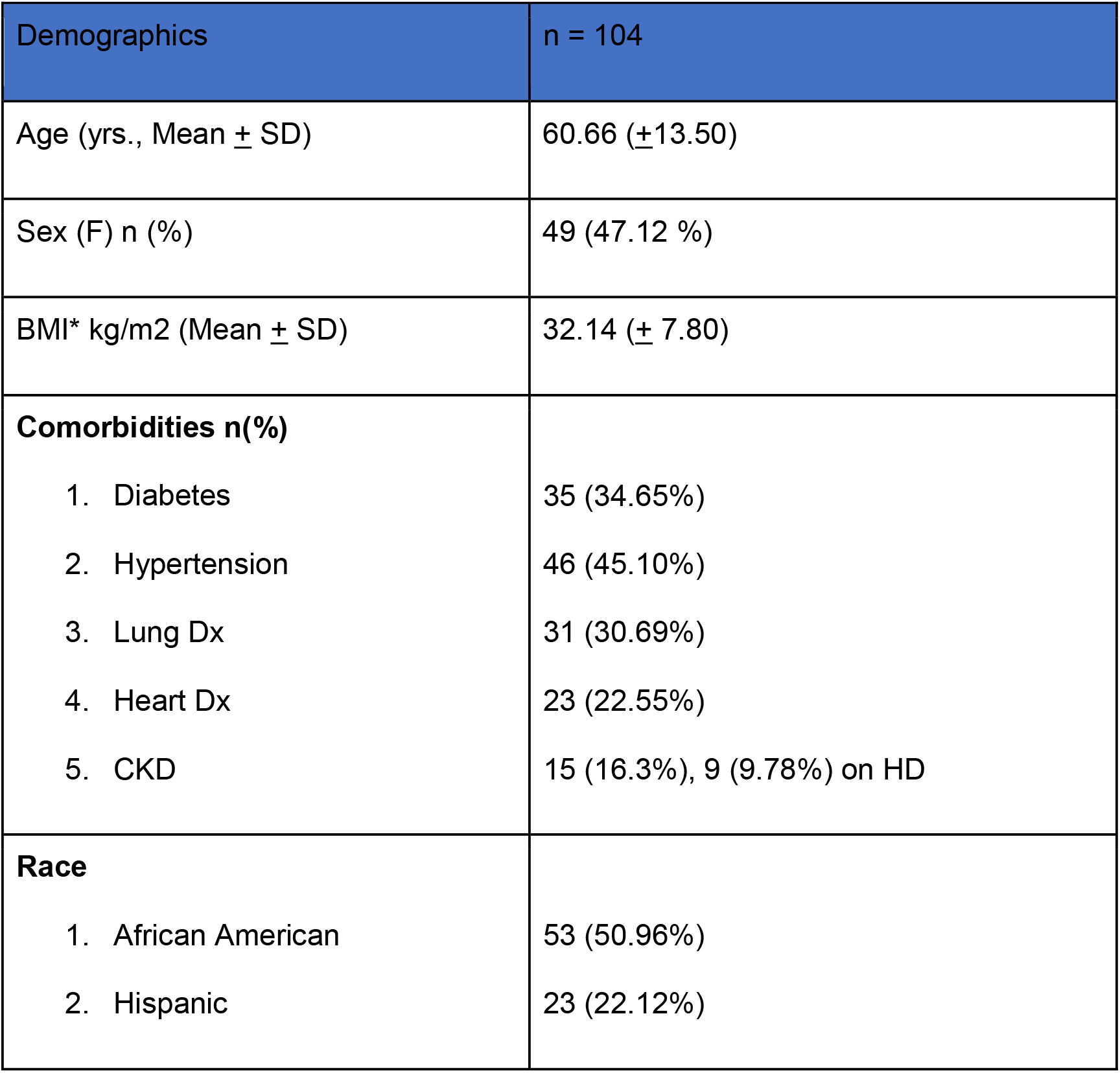
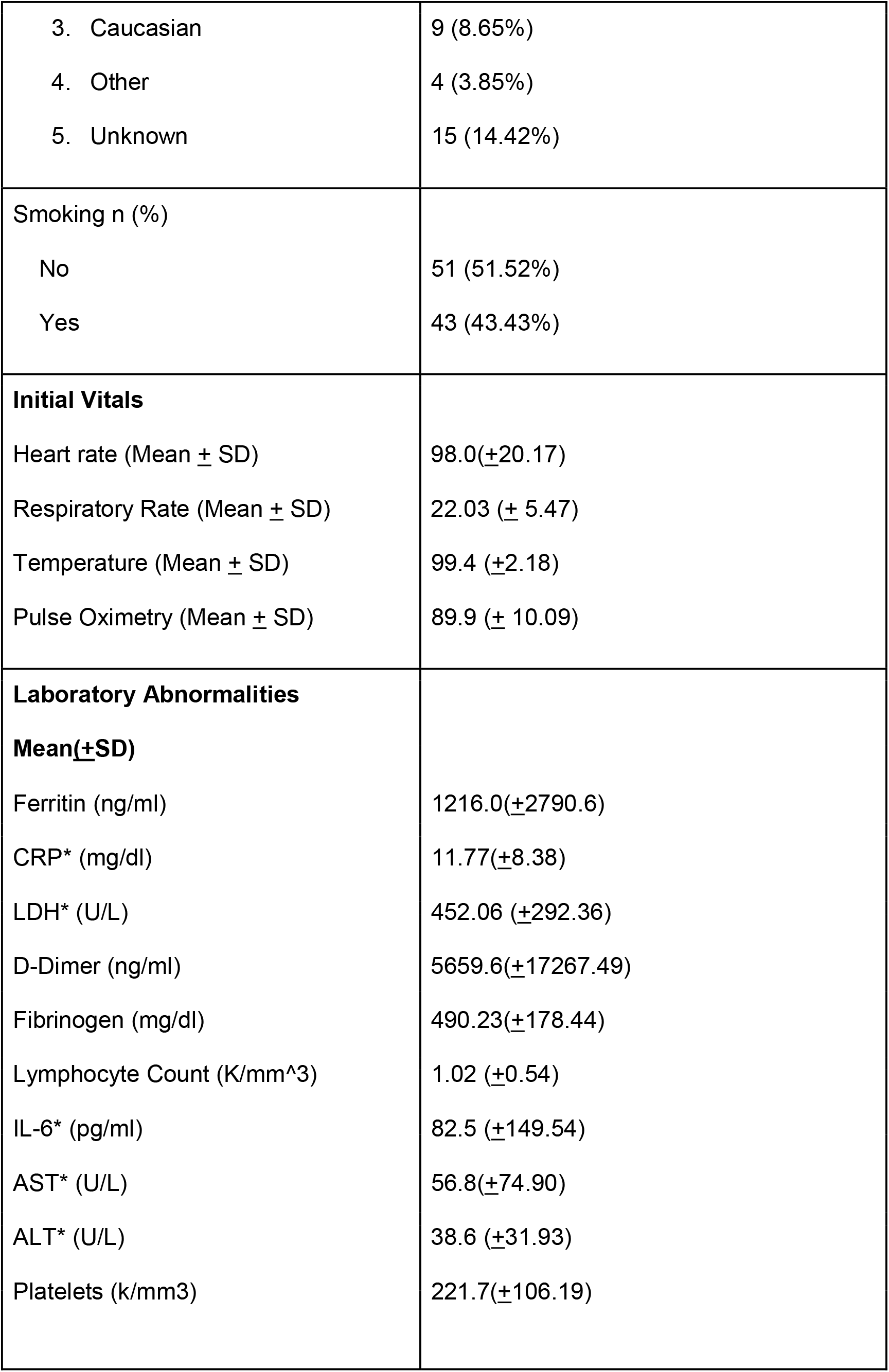
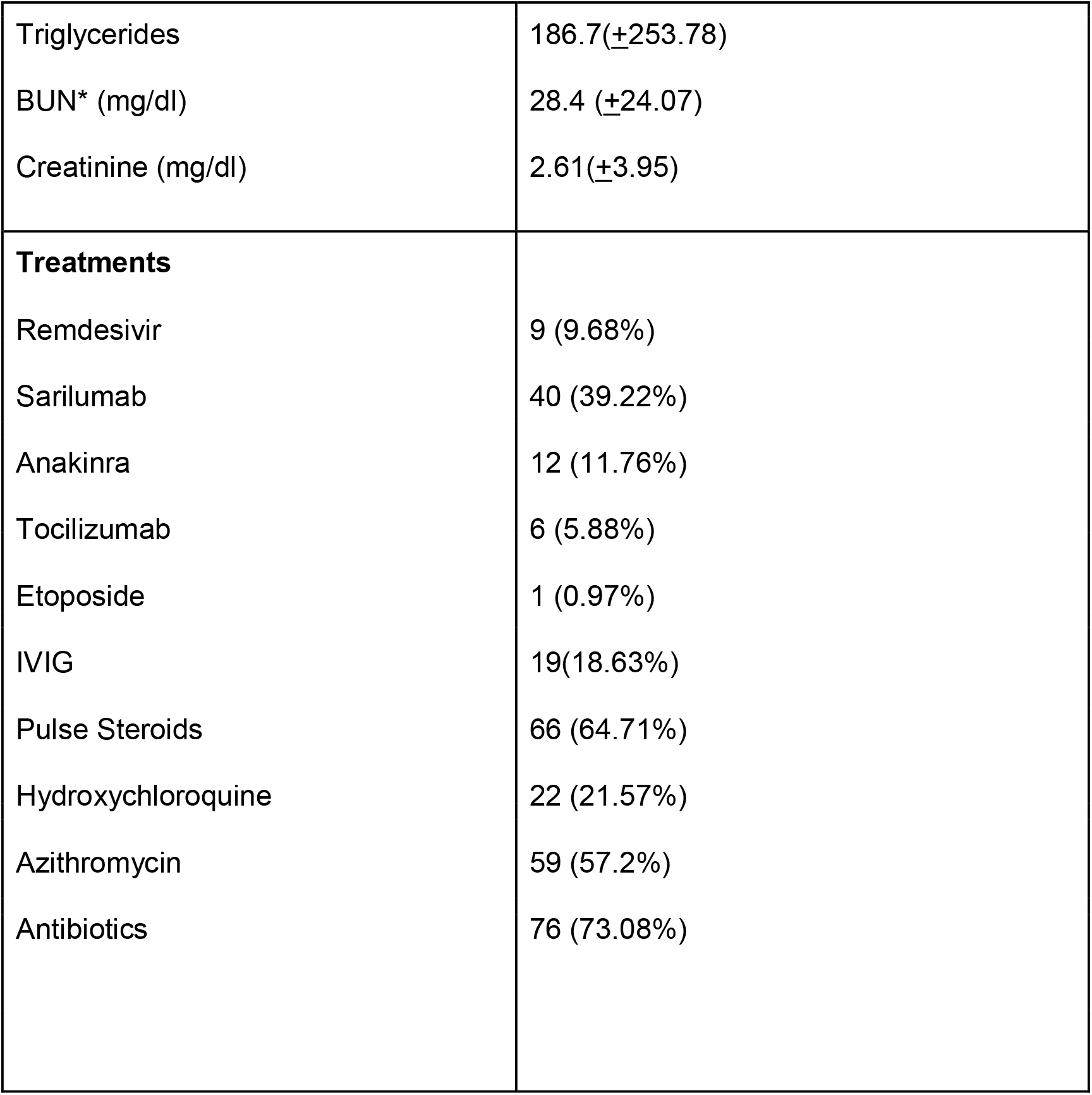
Demographics data including laboratory and clinical parameters.

| Demographics | n = 104 |
| --- | --- |
| Age (yrs., Mean ± SD) | 60.66 (±13.50) |
| Sex (F) n (%) | 49 (47.12 %) |
| BMI* kg/m<sup>2</sup> (Mean ± SD) | 32.14 (± 7.80) |
| **Comorbidities n(%)** |  |
| 1. Diabetes | 35 (34.65%) |
| 2. Hypertension | 46 (45.10%) |
| 3. Lung Dx | 31 (30.69%) |
| 4. Heart Dx | 23 (22.55%) |
| 5. CKD | 15 (16.3%), 9 (9.78%) on HD |
| **Race** |  |
| 1. African American | 53 (50.96%) |
| 2. Hispanic | 23 (22.12%) |
| 3. Caucasian | 9 (8.65%) |
| 4. Other | 4 (3.85%) |
| 5. Unknown | 15 (14.42%) |
| Smoking n (%) |  |
| No | 51 (51.52%) |
| Yes | 43 (43.43%) |
| **Initial Vitals** |  |
| Heart rate (Mean ± SD) | 98.0(±20.17) |
| Respiratory Rate (Mean ± SD) | 22.03 (± 5.47) |
| Temperature (Mean ± SD) | 99.4 (±2.18) |
| Pulse Oximetry (Mean ± SD) | 89.9 (± 10.09) |
| **Laboratory Abnormalities** |  |
| **Mean(±SD)** |  |
| Ferritin (ng/ml) | 1216.0(±2790.6) |
| CRP* (mg/dl) | 11.77(±8.38) |
| LDH* (U/L) | 452.06 (±292.36) |
| D-Dimer (ng/ml) | 5659.6(±17267.49) |
| Fibrinogen (mg/dl) | 490.23(±178.44) |
| Lymphocyte Count (K/mm<sup>3</sup>) | 1.02 (±0.54) |
| IL-6* (pg/ml) | 82.5 (±149.54) |
| AST* (U/L) | 56.8(±74.90) |
| ALT* (U/L) | 38.6 (±31.93) |
| Platelets (k/mm<sup>3</sup>) | 221.7(±106.19) |
| Triglycerides | 186.7(+253.78) |
| BUN* (mg/dl) | 28.4 (±24.07) |
| Creatinine (mg/dl) | 2.61(±3.95) |
| **Treatments** |  |
| Remdesivir | 9 (9.68%) |
| Sarilumab | 40 (39.22%) |
| Anakinra | 12 (11.76%) |
| Tocilizumab | 6 (5.88%) |
| Etoposide | 1 (0.97%) |
| IVIG | 19(18.63%) |
| Pulse Steroids | 66 (64.71%) |
| Hydroxychloroquine | 22 (21.57%) |
| Azithromycin | 59 (57.2%) |
| Antibiotics | 76 (73.08%) |

### High Flow Details

104 (23.3%) of 445 COVID positive patients required HFNT support. Initial HFNT settings were 31.8 (±9.17) L/min of flow, while fractional of inspired oxygen (FiO2) was 90% (±16.7). The average use of HFNT for our population was 4.58 days (±3.28). The minimum settings on HFNT were 10 L flow and FiO2 of 30%, while the maximum settings were 60 L and FiO2 of 100%. Forty-five (43.2%) of patients receiving HFNT progressed to IMV or NPPV. The incidence of hospital associated pneumonia on HFNT was 2.94%. Two patients were excluded from analysis due to short follow-up.

Use of High-Flow for liberation from Mechanical ventilation (IMV+NIPPV)

11 of the IMV patients were successfully extubated to High flow with no re-intubations in this subgroup. Six of the eight patients on NPPV were successfully liberated from NPPV with the use of HFNT.

### Outcomes

The SF ratio significantly improved from 123.5 to 234.5 from day 1 to day 7. Chest X-ray score improved from 18.17 to 16.13 (p < 0.0001), heart rate decreased from 88.2 (±17.13) to 75.7 (±23.13) (p = 0.004) and respiratory rate improved from 29.71 (±18.99) to 26.38 (±16.93) (p = 0.0001) (Table 2). Sixty seven of 104 (64.42%) were able to avoid invasive mechanical ventilation in our cohort. Overall, 45 patients required mechanical ventilation, of which 37 (35.58%) required IMV and 8 patients (7.69 %) required non-invasive positive pressure ventilation (NPPV).

**Table 2:**
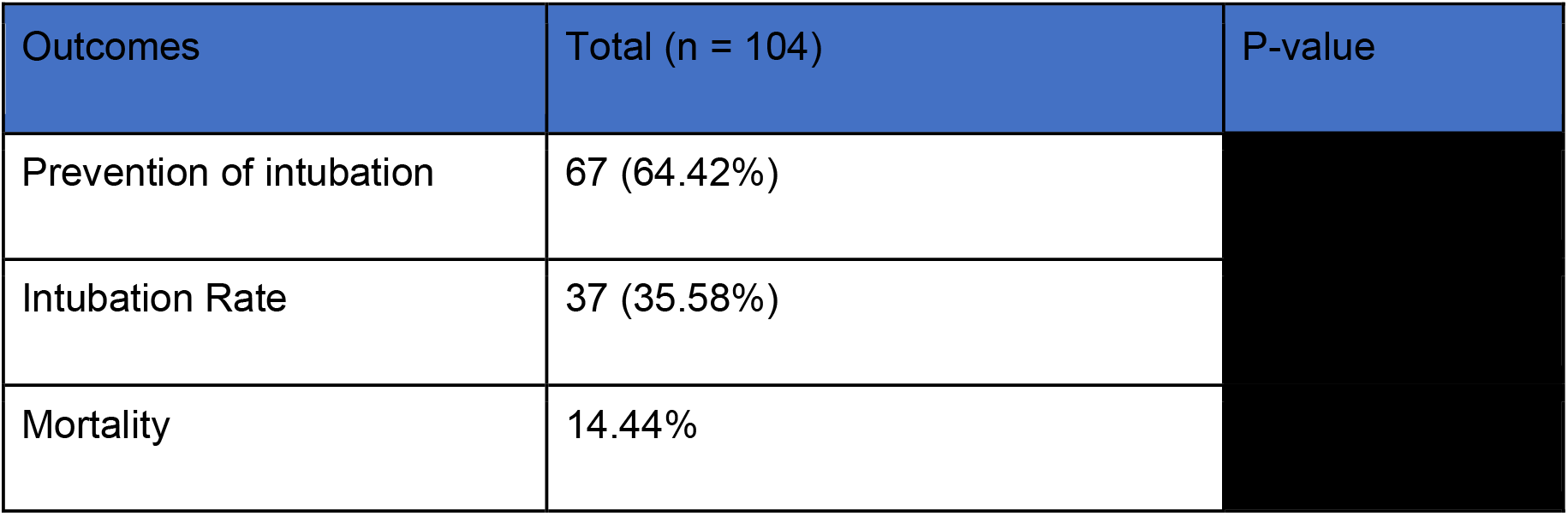
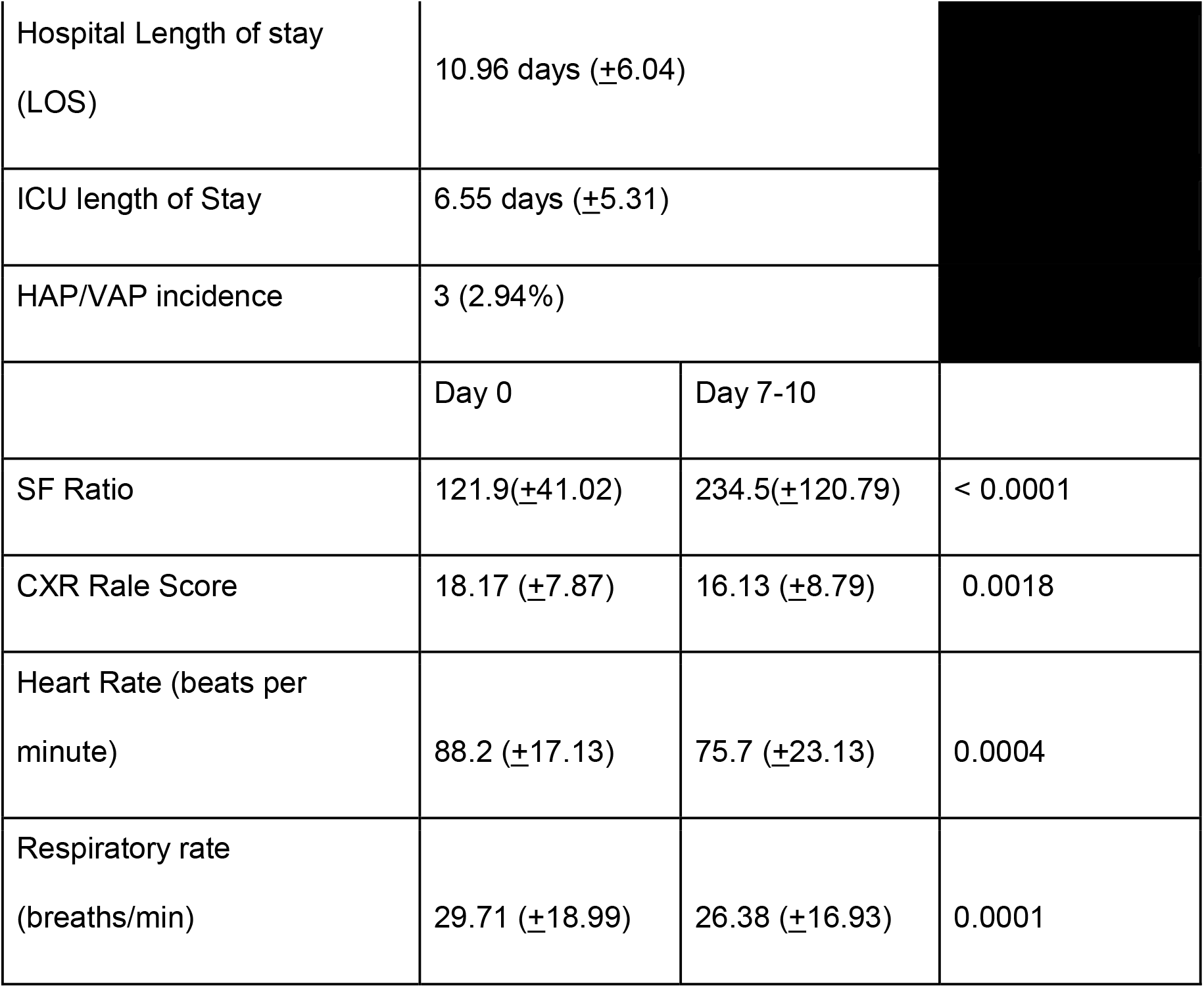
Outcomes of patients treated with HFNT.

Overall, mortality was 14.44% (n=15) in our cohort with 13 (34.4%) in the intubation group and 2 (2.9%) in the non-intubation group. Both the deaths in the non-intubation group were patients transitioned to comfort-directed care. Lastly, 10 of the 13 deaths were related to non-pulmonary organ failure and complications (table 3).

**Table 3:** Clinical descriptors of deceased patients in our COVID19/HFNT cohort.

|  | Age | Comorbidities | BMI | Complications |
| --- | --- | --- | --- | --- |
| Patient 1 | 90 | HTN, Diabetes, PE, HFpEF | 36.4 | Family opted for comfort/Hospice care |
| Patient 2 | 78 | Stage 5 CKD, HTN, Metastatic prostate cancer | 29 | Patient had a prolonged stay on the ventilator with failure to wean off the vent.<br>Acute on Chronic Renal failure requiring Dialysis<br>Family decided to opt for comfort/Hospice. |
| Patient 3 | 70 | OSA, Asthma, ILD,<br>severe PH, HFrEF,<br>Diabetes, HTN, CAD,<br>aortic stenosis | 41 | Heart failure exacerbation<br>Family opted for comfort/Hospice care |
| Patient 4 | 65 | HFrEF with AICD,<br>Afib, Diabetes, HTN,<br>CKD, BRASH<br>syndrome,<br>Congestive hepatic<br>failure | 32.4 | Heart failure exacerbation<br>Renal failure requiring Dialysis<br>Ascites requiring Paracentesis<br>Invasive mechanical ventilation |
| Patient 5 | 75 | Left lung transplant,<br>Severe COPD,<br>Benign pleural<br>asbestosis, chronic<br>hypoxemic respiratory<br>failure | 20.3 | Superimposed klebsiella pneumonia<br>Invasive mechanical ventilation<br>Acute kidney injury |
| Patient 6 | 56 | Chronic hypoxic<br>respiratory failure,<br>COPD, OSA, HTN,<br>seizure disorder | 39.5 | Acute renal failure with hyperkalemia<br>Invasive mechanical ventilation |
| Patient 7 | 76 | HTN, arthritis | 33.6 | Acute kidney injury with metabolic acidosis and<br>hyperkalemia<br>Invasive mechanical ventilation |
|  |  |  |  | Hypercapnic respiratory failure |
| Patient 8 | 46 | ESRD on HD, morbid obesity, HTN, HLD, T2DM, former smoker, Chronic Bronchitis, hepatitis B | 35 | Invasive mechanical ventilation<br>Septic shock from staphylococcus bacteremia<br>Acute pulmonary embolism<br>Heparin induced thrombocytopenia<br>Hemophagocytic Lymphohistiocytosis (HLH) |
| Patient 9 | 59 | HTN<br>Glaucoma | 21.93 | Invasive mechanical ventilation<br>Acute arterial thrombosis of left common iliac artery<br>Compartment syndrome requiring fasciotomy<br>Acute renal failure<br>Non-ST elevation myocardial infarction |
| Patient 10 | 69 | HFpEF, Aortic Stenosis, COPD on home oxygen, Renal transplant with graft failure, ESRD on HD, DVT, CVA, T2DM, HTN, OSA | 28.17 | Pulmonary edema requiring emergent ultrafiltration<br>Invasive Mechanical ventilation |
| Patient 11 | 66 | HTN, HLD, Stage III CKD, current smoker | 27.3 | Invasive Mechanical ventilation<br>Acute Pulmonary embolism<br>Acute kidney injury requiring HD |
| Patient 12 | 67 | Hypertension,CKD<br>Diabetes, Sacral<br>decubitus ulcers | 30.45 | Tracheostomy<br>Invasive Mechanical Ventilation<br>Delirium<br>Ventricular tachycardia<br>Acute renal failure |
| Patient 13 | 73 | Lung transplant,<br>Pulmonary fibrosis,<br>HTN, Atrial fibrillation,<br>CAD with cardiac<br>bypass surgery | 27.08 | Invasive Mechanical Ventilation<br>DVT<br>Acute kidney injury<br>Heparin induced thrombocytopenia |
| Patient 14 | 50 | HFrEF40-45%, Renal<br>Transplant with failed<br>graft, HD, breast<br>cancer s/p<br>mastectomy, DM,<br>HTN | 50 | Septic shock from staphylococcus epidermidis bacteremia<br>Heparin induced thrombocytopenia<br>Cardiac arrhythmias |
| Patient 15 | 41 | bipolar disorder,<br>endometrial<br>adenocarcinoma s/p<br>hysterectomy/salping<br>ectomy | 37.12 | New Renal Cell carcinoma with Metastasis<br>Acute respiratory distress syndrome. |

\*HTN - hypertension, PE- pulmonary embolism, HFpEF- Heart failure with preserved Ejection fraction, CKD - chronic kidney disease, OSA- Obstructive sleep apnea, ILD- interstitial lung disease, PH- pulmonary hypertension, HFrEF - heart failure with reduced Ejection fraction, CAD - coronary artery disease, COPD- chronic obstructive pulmonary disease, ESRD - End stage renal disease, HD- hemodialysis, DVT- deep venous thrombosis, CVA - cerebrovascular accident

As of this writing, 48 patients from the HFNT group were discharged from the hospital with LOS 10.9 days (±6.04). ICU length of stay for the 38 patients discharged from ICU was 6.55 days (±5.31). ICU LOS was higher for the intubation group (10.45 days vs 4.05 days, p=0.0008).

### Intubation versus non-intubation (continued HFNT) group

The average duration of high flow use was higher in the non-intubation group. (5.38 days vs 3.11 days, P=0.0023). There were no statistically significant differences between the intubation and non-intubation groups in terms of demographics (age, sex, BMI, most comorbidities, smoking). Hypertension and smoking prevalence were higher in the intubation group. Amongst laboratory markers, D-dimer, LDH and Fibrinogen was higher in the non-intubation group while ferritin, triglycerides, IL-6, AST, BUN and creatinine were higher in the intubation group (table 4). SF ratios were significantly different between the two groups at baseline, with the intubation group having much lower SF ratios compared to those who remained on HFNT (111.03 vs 127.9, p=004). There was greater improvement in SF ratio and chest X-ray score (Figure 3) in the non-intubation group (Table 5). Patients in the intubation group had higher tocilizumab use, whereas Anakinra, IVIG and antibiotics were more common in the non-intubation group.

**Table 4:**
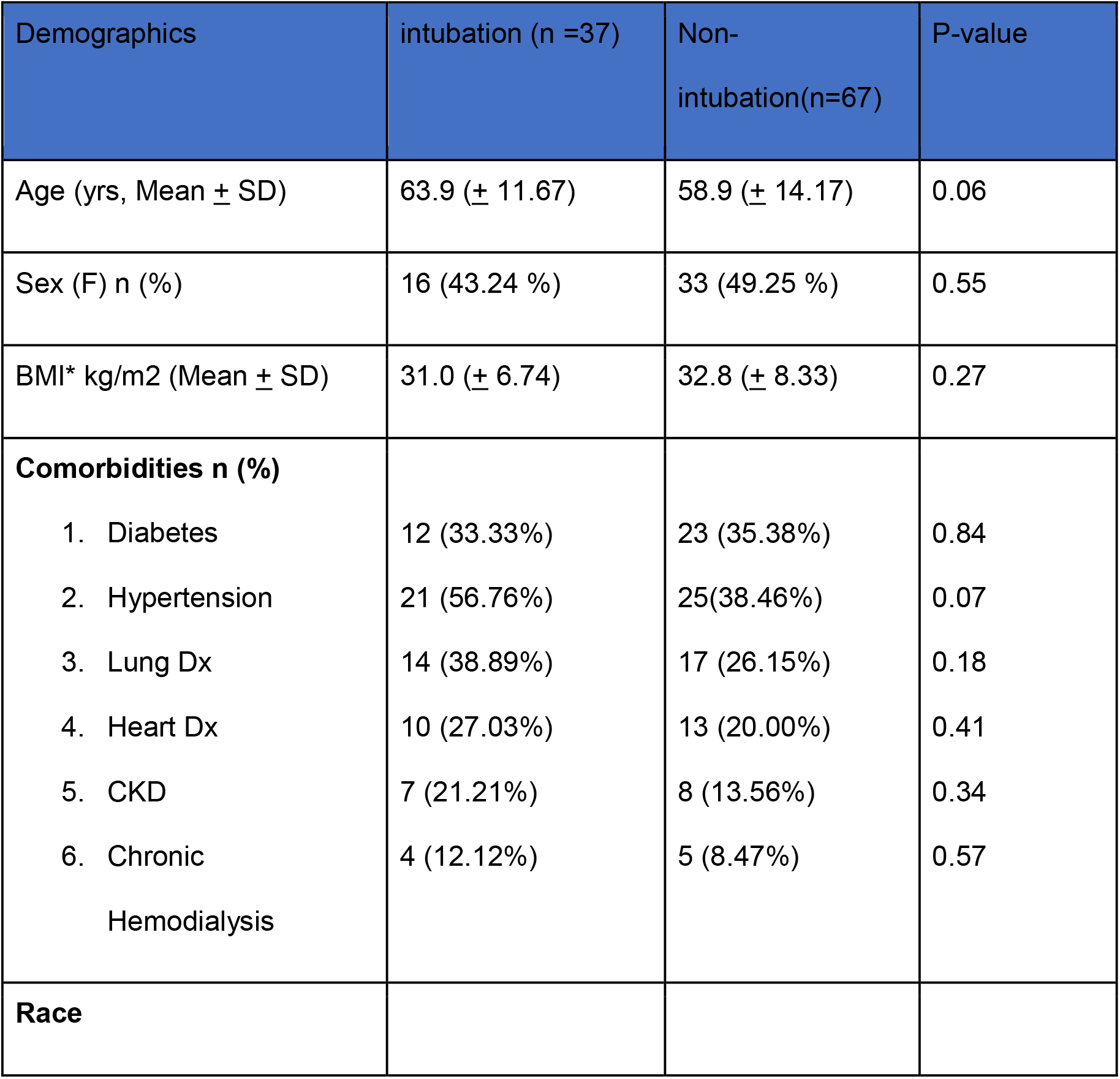
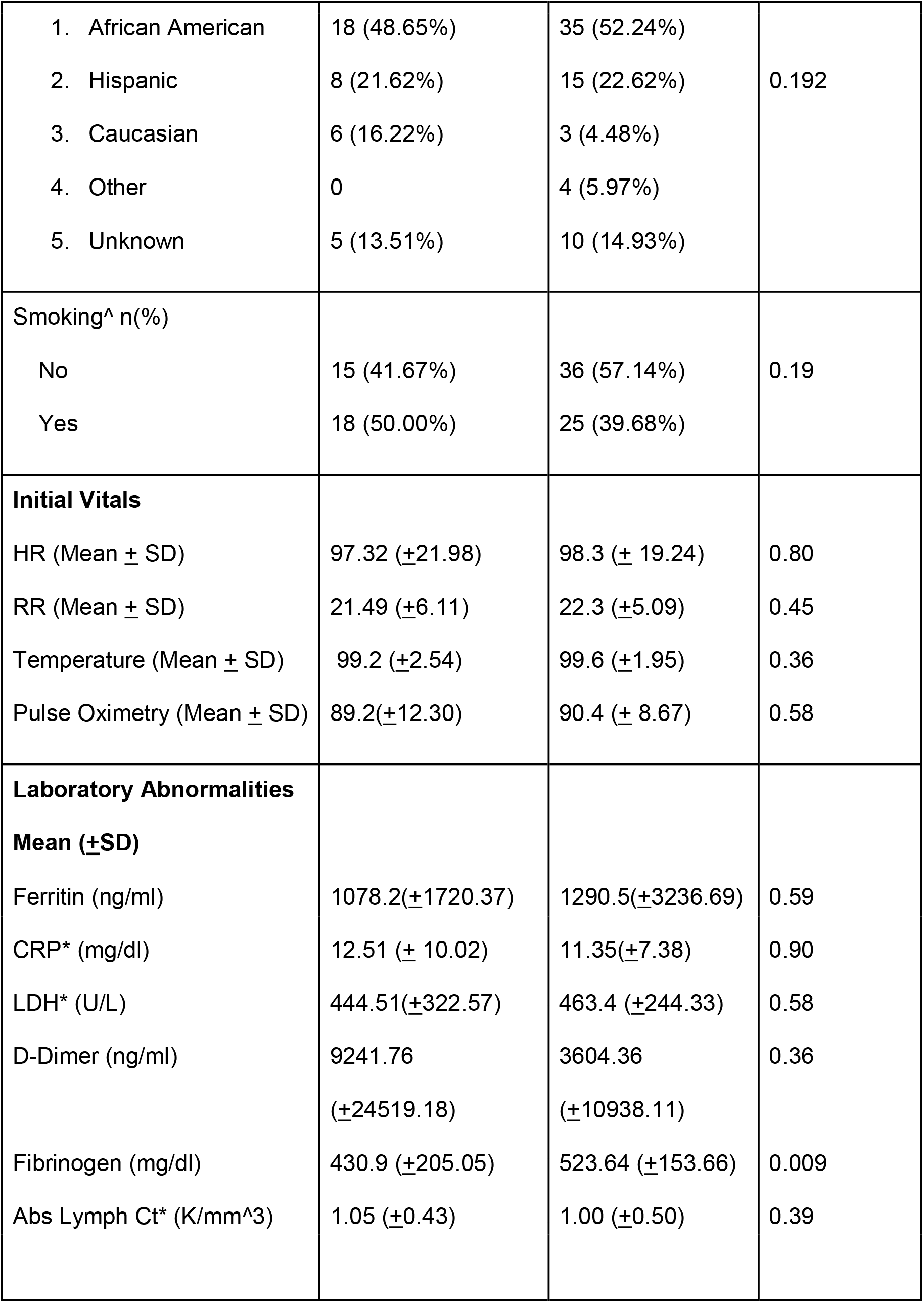
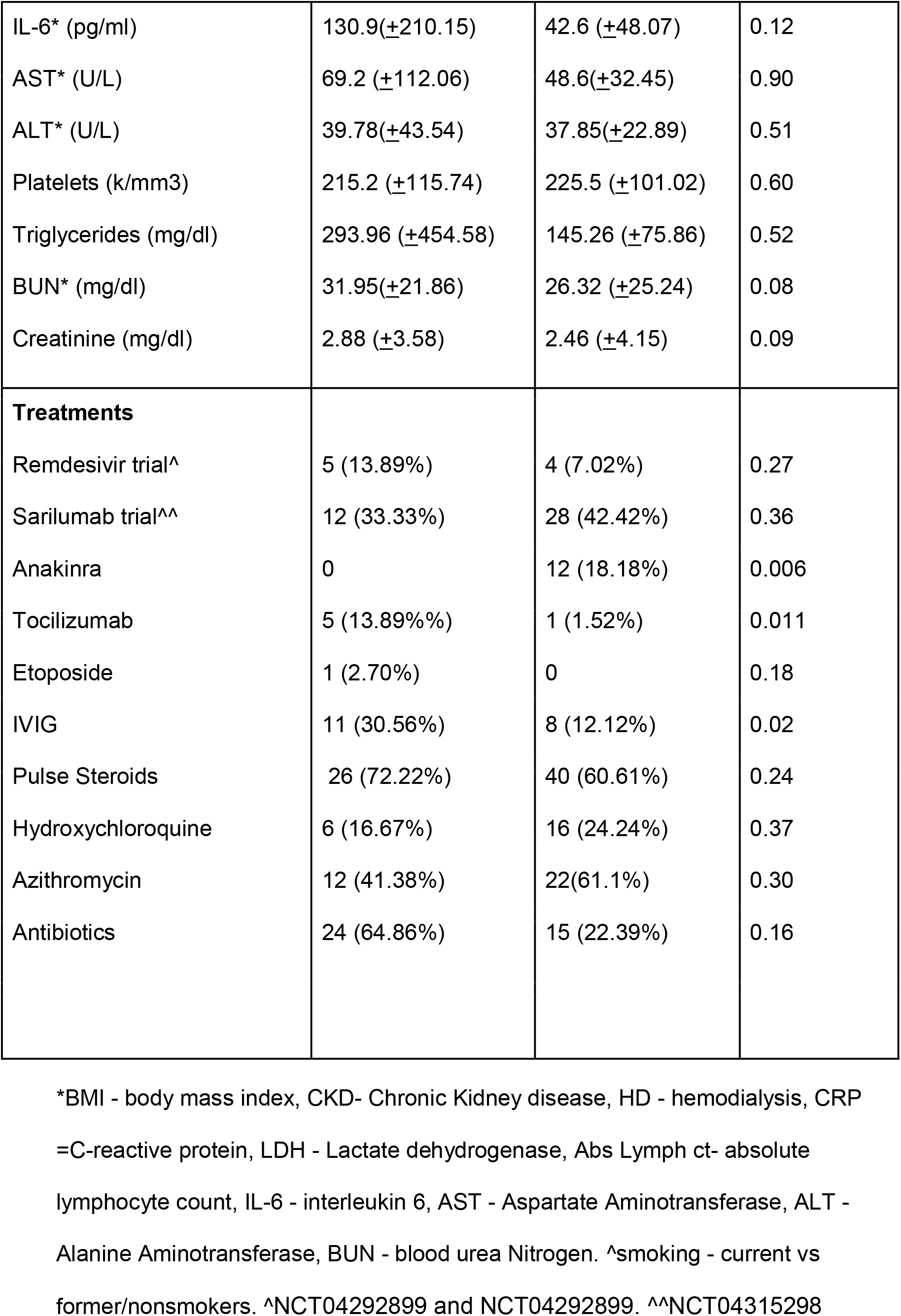
Comparing demographics data between intubation and non-progression groups.

**Figure 3:**
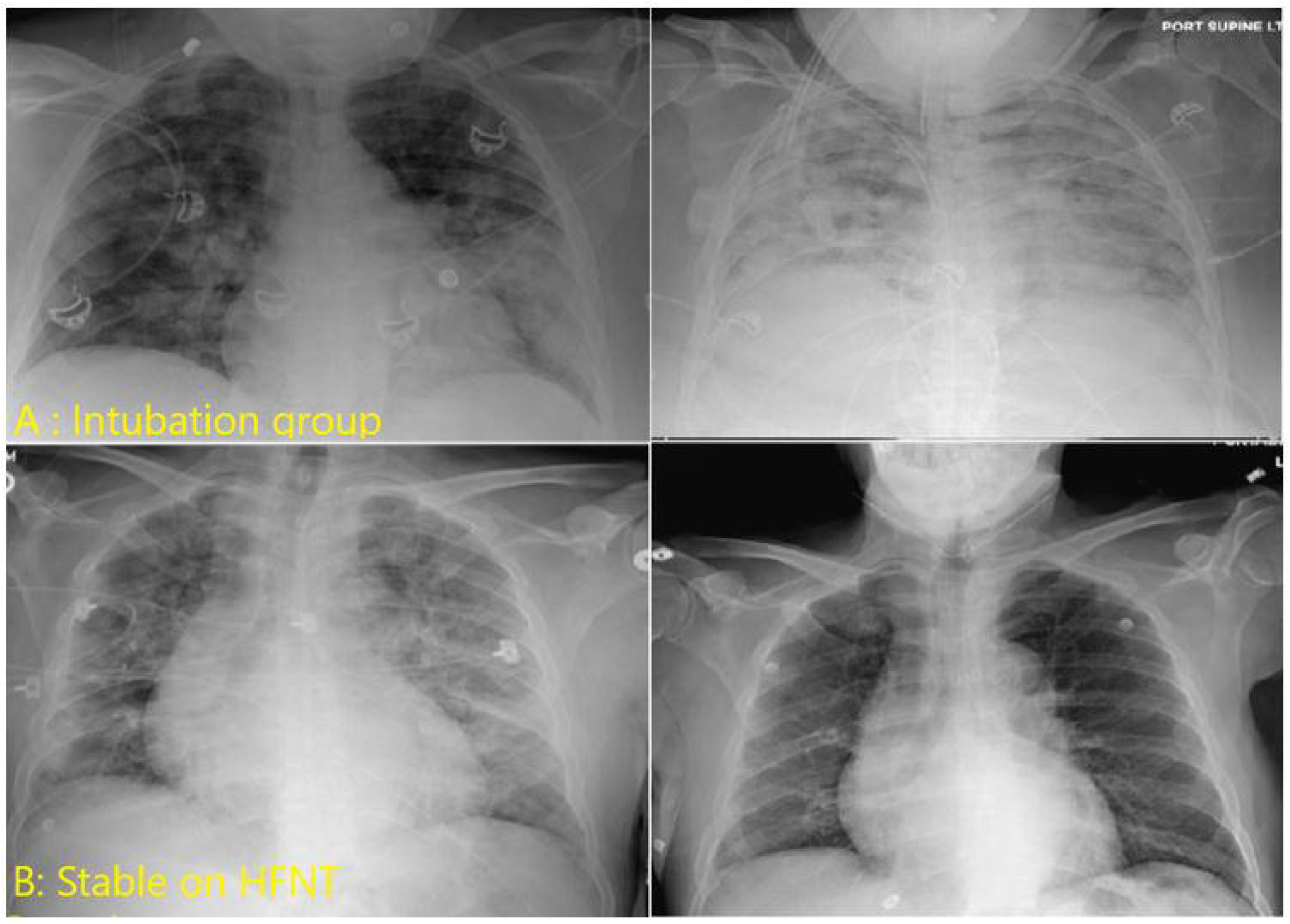
Progression of Chest imaging for patients on High Flow. Image A: Worsening bilateral infiltrates in Intubation group Image B: Non-intubation group, improved infiltrates

**Table 5:** Comparing outcomes between intubation and non-intubation groups.

| Outcomes | Intubation | Non-intubation | P-value |
| --- | --- | --- | --- |
| Mortality | 13 (35.1%) | 2 (2.9%) | 0.0018 |
| Hospital LOS*<br>(days) | 13.67(±7.97) | 9.7(±4.6) | 0.03 |
| ICU LOS<br>(days) | 10.45 (±6.12) | 4.05 (±2.64) | 0.0008 |
| HAP/VAP incidence | 3 (8.57%) | 0 | 0.017 |
| Change in SF Ratio | 40.5 (±67.90) | 141.4 (±117.14) | 0.0001 |
| Change in CXR Rale<br>Score | -0.13(±11.18) | -3.2(±8.50) | 0.09 |
| Change in HR<br>(beats/minute) | - 7.65 (±20.69) | -7.95(±19.19) | 0.94 |
| Change in RR<br>(breaths/min) | -2.32 (±9.32) | -4.4(±8.39) | 0.27 |

\*LOS - Length of stay, HAP/VAP - hospital acquired pneumonia/ventilator associated pneumonia, SF ratio - oxygen saturation/fraction of inspired oxygen, HR- heart rate, RR- respiratory rate

Mortality and incidence of Ventilator associated Pneumonia/Hospital acquired pneumonia was statistically higher in the intubation groups. Figure 4 shows better survival for the non-intubation group compared to the intubation group.

**Figure 4:**
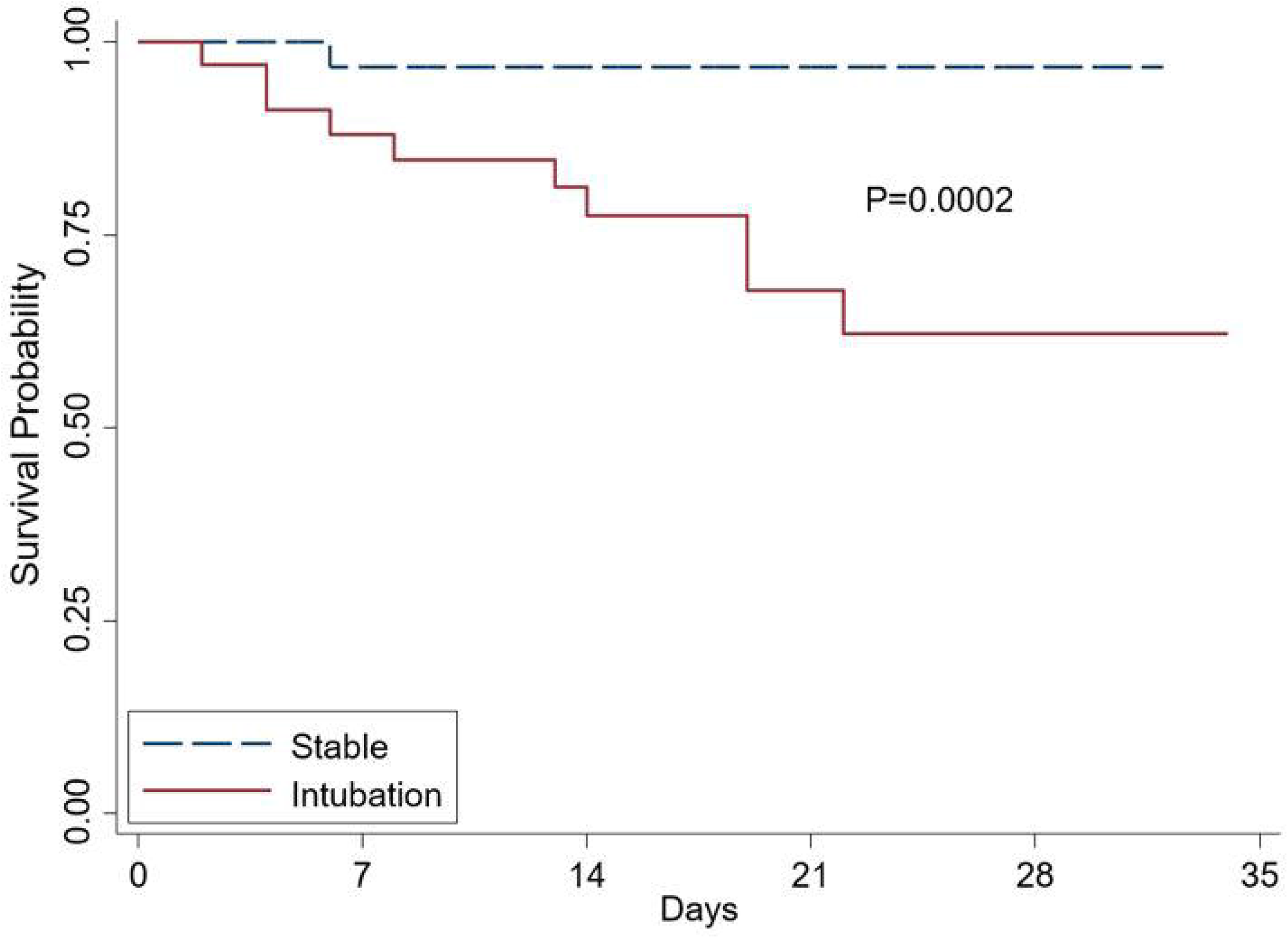
Kaplan Meir estimate of survival of HFNT patients, comparing intubation versus non-intubation (continued HFNT) groups

### Prediction Model

In the univariate analysis, history of hypertension, chronic kidney disease (CKD) or having a composite comorbidity score of 1 or greater was predictive of progression to intubation. In terms of laboratory markers, elevated triglycerides (>300 mg/dl) and lower fibrinogen (<=450) were predictive in univariate analysis. S-F ratio <100 (OR = 2.3) was also a significant predictor in univariate analysis. In the multivariate analysis only S-F ratio (<100), history of chronic kidney disease and Fibrinogen (<450 mg/dl) were predictive of intubation (table 6). Figure 5 shows the ROC curve for our prediction model (ROC = 0.7229)

**Table 6:**
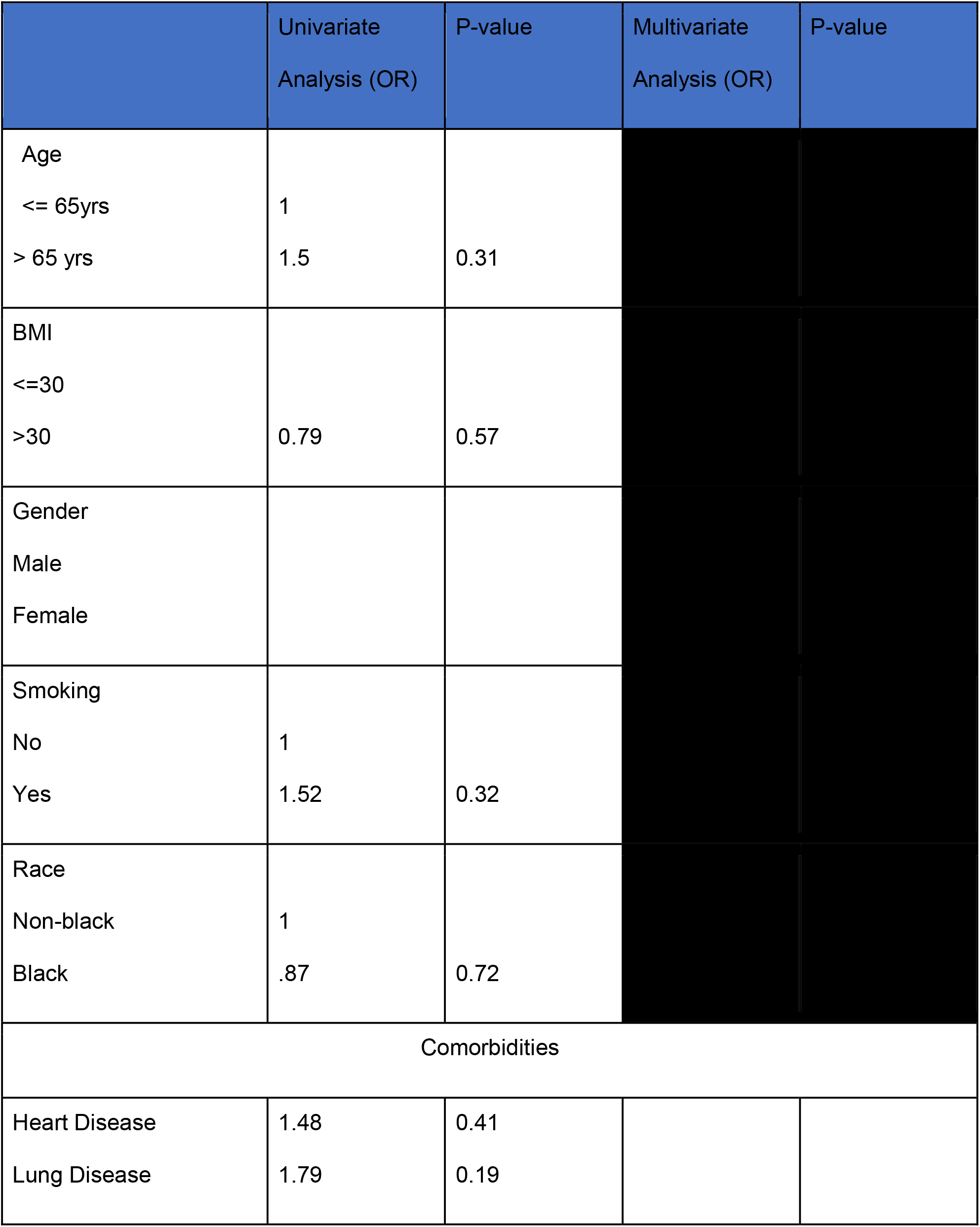
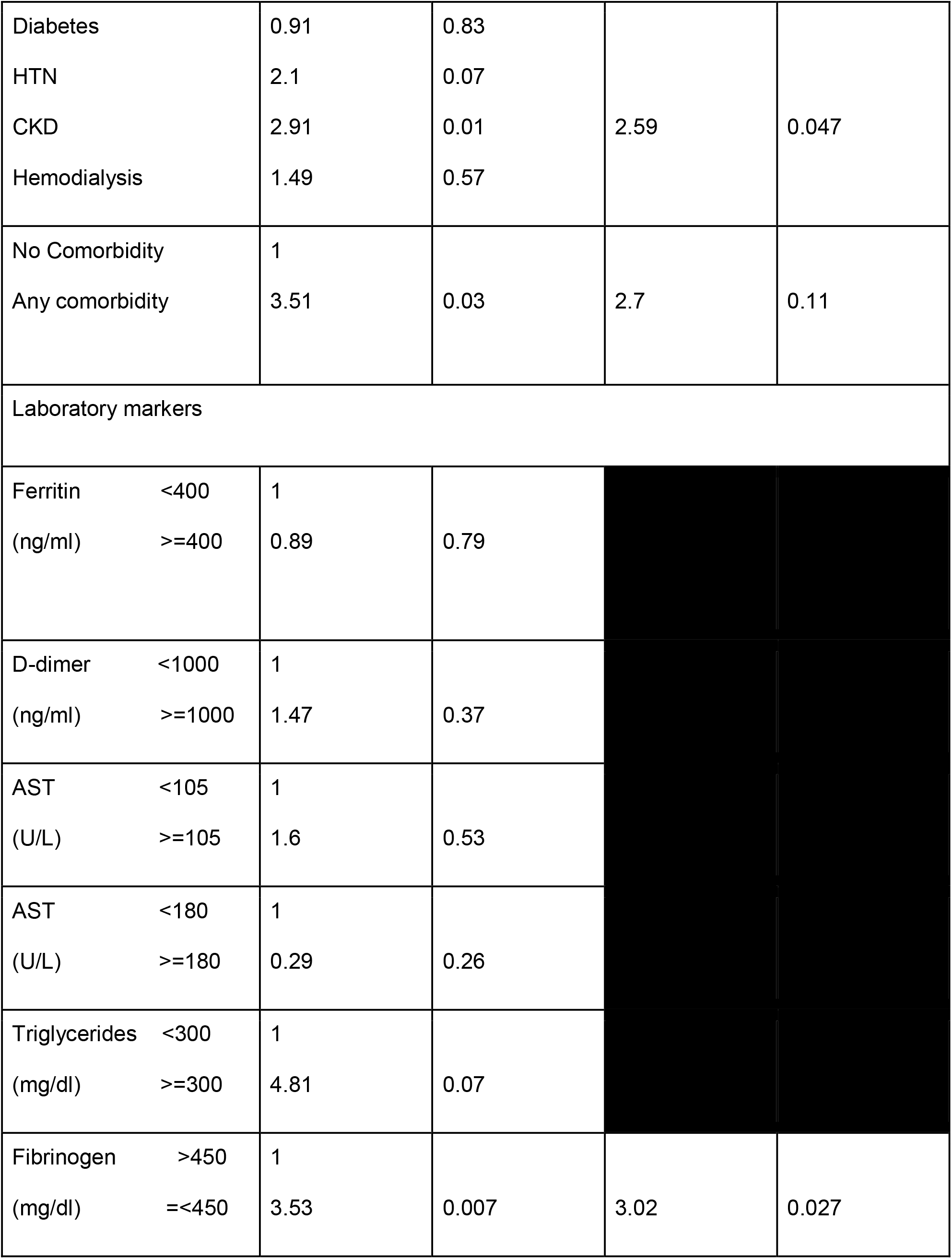
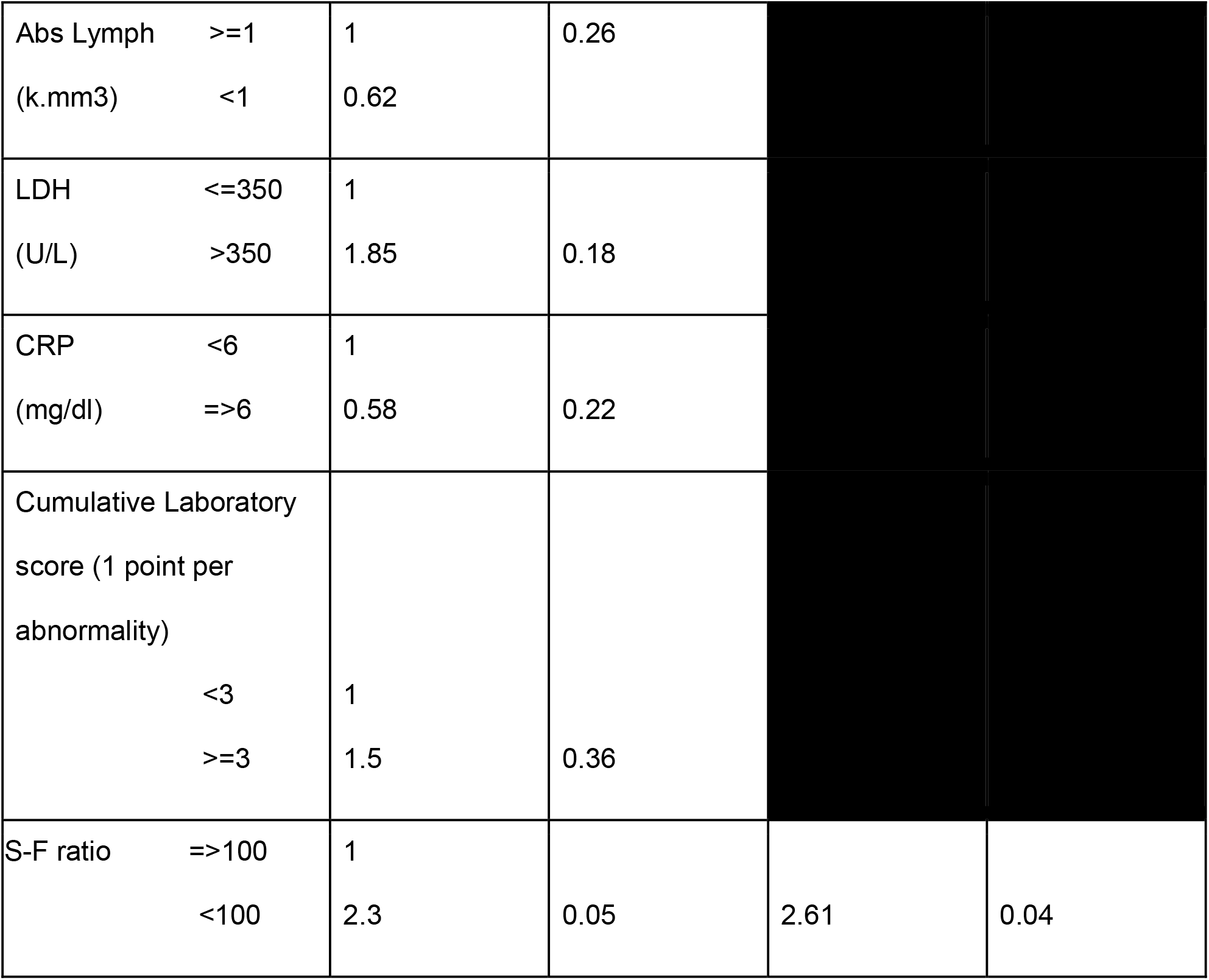
Demographic, Clinical and laboratory predictors of Intubation using Multivariable Logistic regression.

**Figure 5:**
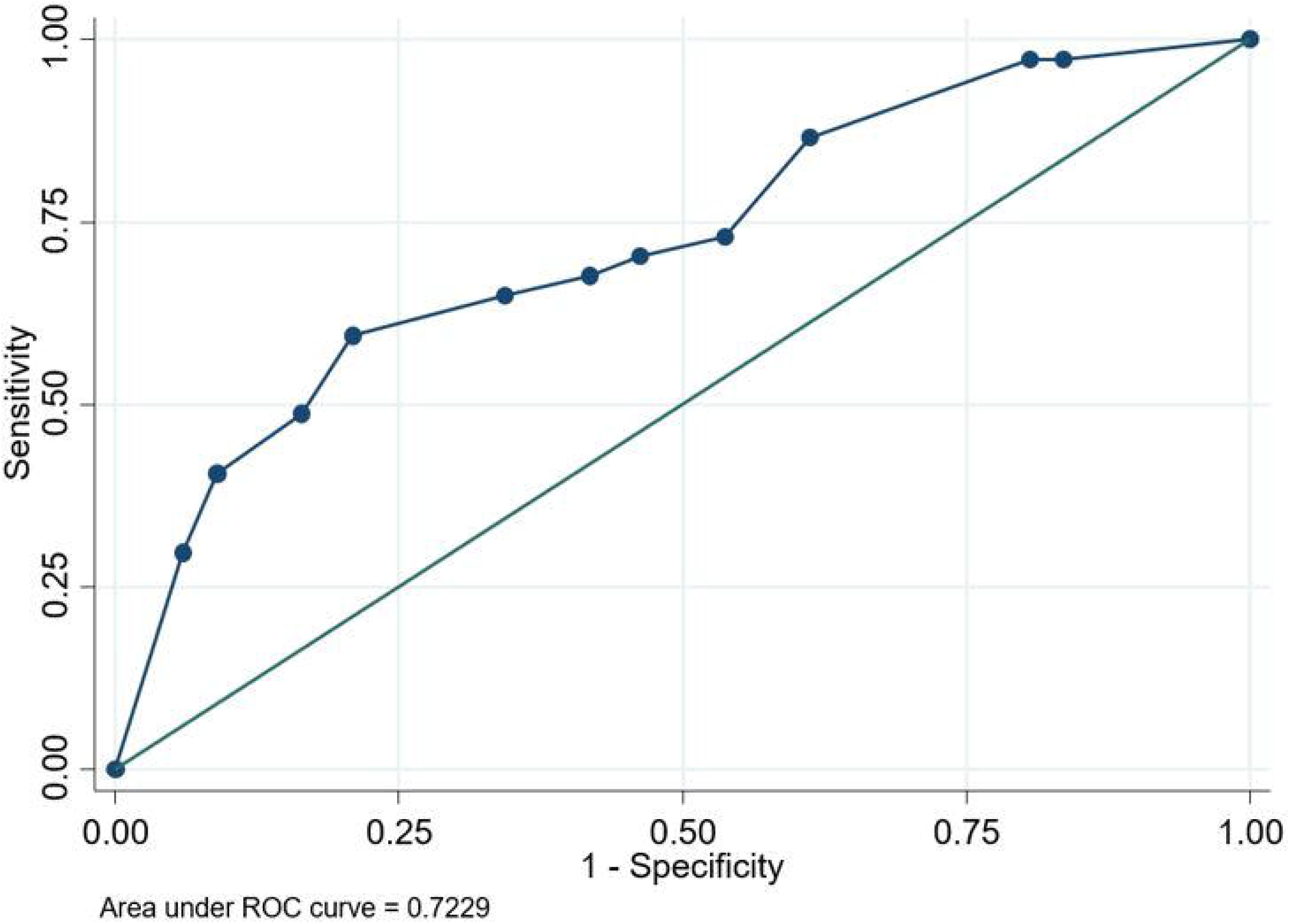
ROC curve of the predictive model for intubation

## Discussion

In this retrospective review of patients with COVID-19 and acute hypoxemic respiratory failure we found that 104 patients (23.3%) were treated initially with HFNT, of which 64.4% remained on HFNT and were able to avoid escalation to non-invasive and invasive mechanical ventilation. The 67 non-intubation patients (continued HFNT therapy) had a significant improvement in oxygenation and reduction in incidence of hospital-acquired pneumonia compared to those who progressed to intubation or NPPV. While the survival advantage cannot be attributed to HFNT based on our study’s retrospective design, use of HFNT did not result in worsened outcomes either. Majority of the patient mortality was attributed to the high burden of comorbidities (metastatic cancer, underlying renal and cardiac conditions, obesity, smoking and bacteremia), rather than progression of respiratory failure on HFNT (Table 5).

In similar patients in Italy and China, the intubation rate has been reported between 70-90%. (3, 20). In addition, our group also had a very high burden of comorbid disease, including underlying lung disease and tobacco use. Among our cohort of patients, 30.69% of patients had underlying lung disease and 43.43% were current smokers. In comparison, early case series reports from China only describe 1.1-3.1 % of patients with chronic obstructive pulmonary disease (COPD) (1, 4, 22), whereas case series from the Lombardy region of Italy reports 4% of patients with COPD (3). Bhatraju et al. reported only one patient with COPD in their recent case series of 21 patients from the Seattle region. (23) The rate of smokers in these studies was also low compared with our group’s prevalence of 43.43%. There was no statistically significant difference in our group between those with and without underlying lung disease with regards to progression to invasive mechanical ventilation. In addition, Hypertension and CKD were also shown to be predictive of intubation in our univariate analysis, with CKD also a predictor in multivariate analysis. Chronic uremia in presence of hypertension leads to chronic left ventricular hypertrophy and other structural changes to the myocardium leaving the patients vulnerable to very small amounts of fluid shifts; subsequently leading to pulmonary edema.(24) CKD has also previously been shown to have worse outcomes including mortality in patients diagnosed with pneumonia.(25) A fibrinogen level of less than 450 mg/dL was found to be predictive of intubation in both univariate and multivariate analysis. fibrinogen is an acute phase reactant and it is possible that patients that present with a fibrinogen <450 mg/dL may be presenting in a later stage of disease and less amenable to antiviral or anti-inflammatory therapies during support with HFNT

Prevention of avoidable invasive mechanical ventilation with HFNT is significant as by nature it avoids incidence of ventilator-associated pneumonia, reduces the need to use medications such as sedatives in which shortages are being reported in the current public health crisis (26, 27). The reported mortality in patients requiring invasive mechanical ventilation in COVID-19 is 90% (3, 4, 20). Our study shows mortality to be much lower when IMV can be avoided. In addition, HFNT can also decrease utilization of ventilators, sedatives in the setting of a global pandemic; thus, representing a viable alternative to IMV.

Gattinoni and colleagues have previously reported high respiratory compliance despite a large shunt fraction (28), proposing that COVID-19 patients fall into two groups. The “Type L” or “non-ARDS Type 1” phenotype have low elastance/high compliance and possible loss of hypoxic vasoconstriction mechanisms and often present with profound hypoxemia and low lung recruitability. The “Type H” or “ARDS Type 2” phenotype has increased pulmonary edema and progression to consolidation and requires traditional management strategies of higher PEEP and lower tidal volumes (29). We have experienced similar patient subgroups in our practice. As HFNT only provides a modest PEEP effect (i.e. 3-5 cmH_2_O at flow rates of 30-50 lpm with mouth closed) (30) patients with predominant Type L physiology who do not require the higher positive pressure benefit from the oxygenation support that HFNC can provide noninvasively. HFNT can lead to a high oxygen reservoir by reducing anatomical dead space in the nasopharynx (31). Furthermore, IMV using high tidal volume (which is often employed in Type L patients) has shown to have inflammatory cytokine release in ARDS patients, including IL-6, both in critically ill humans (32, 33) and murine models (34, 35); IL-6 in particular is one of the pathologic mechanisms for lung injury in COVID-19 (36, 37). Thus, use of HFNT should be a priority in patients with severe COVID-19 respiratory failure.

We elected to use SF ratio than traditional PF ratios in this study for several reasons. SF ratios have been well correlated to standard PF ratios in adult and pediatric populations (38, 39). SF ratios < 235 predict moderate-severe respiratory failure with 85% specificity (39). Our cohort overall showed moderate to severe hypoxemic respiratory failure (mean SF ratio 123 overall), but nonetheless ~64.4 % of our cohort could still be supported with high flow oxygen alone. In contrast, Wang et.al showed only 37% of COVID-19 patients did not progress on HFNT when the P-F ratio was less than 200(40). Additionally, lab draws, and arterial blood gases were limited during a pandemic to minimize staff exposure when possible. Hence, ABGs were not routinely collected as part of standard clinical practice at our institution

There has been debate worldwide about the use of HNFT or other methods of non-invasive ventilation out of concerns for increased disease transmission. During the 2003 SARS outbreak, hospital workers had development of SARS in only 8% of HFNT patients. (41) Studies have not shown that bacterial environmental contamination was increased in the setting of HFNT use (13, 14, 42). An in-vitro study mimicking clinical scenarios including HFNT with mannequins only revealed proximal dispersion of secretions to the face and nasal cannula itself.(43, 44) A recent study with healthy volunteers wearing high-flow nasal cannulas at both 30 L/min and 60 L/min of gas flow did not report variable aerosolization of particles between 10-10,000 nm, regardless of coughing, when compared with patients on room air or oxygen via regular nasal cannula.(45)

This study has several limitations. First, it was retrospective in nature as developing a prospective trial on the initial management of acute hypoxemic respiratory failure in the face of an evolving public health crisis is difficult. Second, we could not reasonably analyze a control arm as our endpoint was prevention of mechanical ventilation. Developing a prospective study during a pandemic situation is impractical without first determining clinical equipoise. Third, we do not report on arterial pH or partial pressure of carbon dioxide (PaCO_2_) as many patients did not have baseline or follow up arterial blood gas measurements prior to initiation of HFNT. We recognize that in many clinical trials an elevated PaCO2 was an exclusion criterion for enrollment. (7, 10) Fourth, our data on hospital length of stay was limited since several patients were still hospitalized at the time data was collected.

Institutions around the world have been skeptical about the use of HFNT in CVOID-19 patients. However, based on our findings, we conclude that there is a role for high flow nasal therapy in patients with COVID-19 related severe respiratory failure especially the L-phenotype. Use of HFNT can not only reduce intubation rates, but also has the potential to reduce mortality and morbidity associated with it.

## Data Availability

All data will be available upon request. All requests can be made to the corresponding author.

